# BCG Vaccination Policy, Natural Boosting and Pediatric Brain and CNS Tumor Incidences

**DOI:** 10.1101/2023.04.10.23288314

**Authors:** Samer Singh, Amita Diwakar, Rakesh K. Singh

## Abstract

Bacille Calmette-Guérin (BCG) vaccination supposedly imparts and augments “trained immunity” that cross-protects against multiple unrelated pathogens and enhances general immune surveillance. Gradual reductions in tuberculosis burden over the last 3-5 decades have resulted in the withdrawal of BCG vaccination mandates from developed industrialized countries while reducing to a single neonatal shot in the rest. Concurrently, a steady increase in early childhood Brain and CNS (BCNS) tumors has occurred. Though immunological causes of pediatric BCNS cancer are suspected, the identification of a causal protective variable with an intervention potential has remained elusive. An examination of the countries with contrasting vaccination policies indicates significantly lower BCNS cancer incidence (per hundred-thousand) in countries following neonatal BCG inoculations (n=146) *vs*. non-BCG countries (n=33) [Mean: 1.26 *vs*. 2.64; Median: 0.985 *vs*. 2.8; IQR: 0.31–2.0 *vs*. 2.4–3.2; *P*=<0.0001(two-tailed)]. Remarkably, natural *Mycobacterium* spp. exposure likelihood is negatively correlated with pediatric BCNS cancer incidences in all affected countries [*r*(154): —0.6085, *P*=<0.0001]. Seemingly, neonatal BCG vaccination and natural “boosting” are associated with a 15-20-fold lower BCNS cancer incidence. We attempt to synthesize existing evidence implying the immunological basis of early childhood BCNS cancer incidences and briefly indicate possible causes that could have precluded objective analysis of the existing data in the past. A comprehensive evaluation of immune training as a potential protective variable through well-designed controlled clinical trials or registry-based studies as feasible may be warranted for its potential applications in reducing childhood BCNS cancer incidences.

**Statement of Significance:** Potential causal protective variable for childhood Brain and other CNS (BCNS) tumors has eluded discovery. Neonatal BCG vaccination and boosting by *Mycobacterium tuberculosis* complex exposure seem associated with over 15-20 times lower BCNS cancer incidences. Data suggests neonatal BCG vaccination followed by “boosting” may be preventive for early childhood BCNS cancer incidences.

## INTRODUCTION

Bacille Calmette-Guérin (BCG), a derivative of *Mycobacterium bovis* (a member of *M. tuberculosis* complex), is the most widely used early childhood vaccine that is in use for over 100 years for protecting against tuberculosis (TB) [1]. It offers the greatest protection against miliary TB and tuberculous meningitis and to a lesser extent against pulmonary TB [2]. However, the non-specific protection offered by BCG against other common pathogens, sepsis, and unrelated conditions has been associated with a reduction in early childhood mortality rate by up to 50% in different studies [3-7]. BCG is supposed to provide this protection through the induction of granulopoiesis, the activation of heterologous T-cell immunity, and enhanced non-specific innate immunity committed through epigenetic and metabolic reprogramming [REFs 8-15]. Mechanistically, the inoculation is supposed to bring about functional reprogramming of the cells of innate immune response (*e*.*g*., Monocytes, macrophages, NK cells, etc.) that leads to better immune surveillance and response [REFs 9-16]. In the case of children growing up in ‘hygienic’ conditions, it has been hypothesized to provide the necessary immune stimulus required for a ‘normal’ immune system development that may reduce incidences of common childhood cancers and immune conditions including autoimmune diseases [REFs17-21]. The ability of BCG to potentiate cell-mediated response, which is believed to be important for cancer, has led to its evaluation for preventive and therapeutic potential in many observational studies and clinical trials [REFs 15,18-20,22-28]. However, except in the case of bladder cancer, its preventive and therapeutic potential remains debated and largely uncertain due to the lack of comprehensive and consistent data along with a theoretical framework that could at least attempt to explain different conflicting observations [REFs 15,19,20,22]. The evidence generated so far seems to be overwhelmingly negating the potential of BCG in protecting against cancers. We envision this apparent uncertainty about BCG vaccinations’ potential benefits and conflicting outcomes in different studies to be resulting from the inappropriate generalization of the potency of different BCG vaccines in activating the innate immune system [29,30], omissions about the requirement of boosters [REFs 31-32], longevity of conferred non-specific ‘trained immunity’ that seldom lasts few years in the absence of a booster [31,32,33], treating all cancers as a homogeneous lot disregarding their origin (*e*.*g*., embryonal, mutational: sporadic, germline, primary, secondary, *etc*.), and associated inherent differences, non-consideration of the immune status of subjects and their exposure to a specific intervention and risk factors, *etc*. in different studies reported in [10,15,17-20,22].

Early childhood cancers are hypothesized to result from embryonic aberrant remnants that could have been otherwise eliminated during ‘normal’ development had the appropriate immune stimulus and training been available in the form of common pathogens and microbes to which we have been exposed during evolution [17-20,22,28]. Based on the observation of children brought up in supposedly more hygienic conditions, a critical role for natural exposure to pathogens for immune reprogramming and maturation has been proposed [17-20,34-36]. ‘Hygienic’ conditions have been associated with increased allergy and atopic conditions, susceptibility to life-threatening infections, and even a higher incidence of cancer [REFs.9,11,17,19-22]. However, for the lack of consistent evidence pinpointing a preventive/protective agent or intervention that may have a direct causal relationship, ambivalence about their actual potential in cancer incidence prevention has remained [REFs15,17,19, 22,28].

## RESULTS

### Changing Incidences of Early Childhood Brain and Central Nervous System (BCNS) Cancer and Potential Risk Factors

Brain and other central nervous system (BCNS) cancer remains the second most frequently occurring childhood cancer and the most frequent cause of cancer mortality [37]. The updated estimates for all cancers and ages periodically produced by the International Agency for Research on Cancer, WHO as a part of the GLOBOCAN project (https://gco.iarc.fr/today/home) are a comprehensive source of cancer incidence estimates and related information. Incidentally, the industrialized Western world with high living standards and hygiene has higher incidence rates of both early childhood and later-age BCNS cancer [37]. Most of the countries for which estimates are available have been steadily registering an annual increase in childhood BCNS cancer incidences for the last 3-5 decades [37]. The improvement in TB prevalence and incidences resulting from improved hygiene and living standards globally had also seen a concomitant reversal of BCG mandates, and changes in BCG booster schedules, or altogether scrapping in several countries [38,39]. For the majority of countries/territories with data, the estimated annual % change in total BCNS cancer incidences has been positive (10 out of 42) for recent decades (Supplementary Figure 1) [37]. The childhood BCNS cancer incidence is highest in 0-4-Year-olds (0-4Y-Old or younger than 5 years) and decreases in older 5-14Y-old children. GLOBOCAN study estimates BCNS cancer incidence and associated mortality among 0-4Y-olds during the year 2020 to be 9252 and 4256, respectively, which accounted for about 16% of incidence and 20% of associated deaths from all cancer types combined [37]. Despite, advances in diagnostics and therapeutics, the preventive vaccines and causative risk factors for the majority of early childhood BCNS cancers remain unidentified and uncharacterized [40-43]. Though previously the occurrence of malignant and embryonic origin BCNS tumors in children <5 years has been associated with risk factors like socioeconomic position [41,42,44], allergy [REFs. 42,45,46], infections or exposure to pathogens early in life [REFs. 47,48], human developmental index (HDI), etc. [REFs. 41-48], the identification of potential protective risk factors with an intervention potential (*i*.*e*., employable or actionable) to reduce the incidence of BCNS cancers has so far remained an unfulfilled aspiration.

Compared to BCNS cancers arising in any other age group, the early childhood incidences are predominantly of embryonal origin and malignant, and their incidence rates decrease with age [REFs 40-43]. The scientific community may be aware of the fact that although globally BCNS cancer incidence rates pick up again around 20 years of age and keep going up with age [37,40,41], they are overwhelmingly not of the same origin (up to 10-fold) [42,43]. This offers an opportunity to evaluate any potential role of immune modulation and maturation, more specifically that of BCG vaccination, the most widely given vaccine, on the incidence of childhood BCNS cancer, if any. We surmise that if early childhood BCG vaccination-induced/primed non-specific trained immunity could play any role in cancer incidences arising from abnormal immune system development and maturation, their effect would be more discernible in early childhood cancers, which allegedly arise from embryonic remnants that could not be eliminated during early development for the lack of appropriate and timely immune system stimulation (priming and maturation). The policy on BCG vaccination varies by country (Table 1) [39]. Currently, the majority of middle-to low-income countries follow the BCG vaccination policy for newborns, while non-BCG mandating countries (No-BCG) are mostly affluent high-income countries [49] with very high (vh-) to high (h-) HDI – a composite measure of “a long and healthy life, access to knowledge and a decent standard of living” [50]. The existence of disparities in BCG vaccination policies [39] and TB incidence rates (*i*.*e*., *M. tuberculosis* complex exposure possibility) across countries [51] provides a unique opportunity to test the hypothesis that BCG vaccination and/or “boosting” may be potentially protective (alternatively non-protective) in early childhood BCNS cancer.

**Table 1.** Early Childhood Brain and other Central Nervous System (BCNS) cancer incidence in countries with varying tuberculosis (TB) incidence rates (potential exposure) and correlation analysis

| BCNS Cancer Incidence (BCNS Can. Inc.) in 0-4Y olds (0-4Y) and TB incidence (TB Inc.) rates per 100,000 (100k) for Countries, 2020 |  |  |  |  |  |  |  |  |
| --- | --- | --- | --- | --- | --- | --- | --- | --- |
| Country/Territory [with 'No BCG' for all policy (NB) [39]; <sup>a</sup>HDI high (h-HDI) and Very High (vh-HDI) [50] | <sup>b</sup>BCNS Can. Inc. per 100k (0-4Y) [37] | <sup>c</sup>TB Inc. per 100k [51] | Country/Territory | BCNS Can. Inc. per 100k (0-4Y) | TB Inc. per 100k | Country/Territory | BCNS Can. Inc. per 100k (0-4Y) | TB Inc. per 100k |
| Montenegro (vh-HDI) | 5.4 | 15 | Georgia (vh-HDI) | 2.2 | 69 | Malawi | 0.55 | 138 |
| Estonia (vh-HDI) | 4.3 | 10 | Iran (h-HDI) | 2.2 | 12 | Myanmar | 0.55 | 311 |
| United States (NB; vh-HDI) | 4.3 | 2.3 | Peru (h-HDI) | 2.2 | 117 | Guatemala | 0.53 | 25 |
| Greece (NB; vh-HDI) | 4.2 | 4.2 | Syria | 2.2 | 18 | Mongolia (h-HDI) | 0.53 | 428 |
| Canada (NB; vh-HDI) | 4.1 | 5.4 | Colombia (h-HDI) | 2 | 34 | Uganda | 0.51 | 199 |
| Israel (NB; vh-HDI) | 4 | 2.1 | Iraq | 2 | 26 | Mauritania | 0.43 | 85 |
| Cape Verde | 3.9 | 39 | Oman (vh-HDI) | 2 | 7.9 | Mozambique | 0.43 | 361 |
| Slovakia (NB; vh-HDI) | 3.9 | 3.2 | United Arab Emirates (vh-HDI) | 2 | 0.77 | Rwanda | 0.42 | 57 |
| Slovenia (NB; vh-HDI) | 3.9 | 4.1 | Venezuela (h-HDI) | 2 | 44 | South Sudan | 0.41 | 227 |
| Croatia (vh-HDI) | 3.8 | 5 | Morocco | 1.9 | 96 | Eritrea | 0.4 | 80 |
| Suriname (NB; h-HDI) | 3.8 | 29 | Singapore (vh-HDI) | 1.9 | 46 | Lesotho | 0.39 | 592 |
| North Macedonia (h-HDI) | 3.6 | 12 | Libya (h-HDI) | 1.8 | 59 | Mali | 0.36 | 51 |
| Belarus (vh-HDI) | 3.5 | 28 | China (h-HDI) | 1.7 | 57 | Burundi | 0.34 | 103 |
| Latvia (vh-HDI) | 3.5 | 20 | El Salvador | 1.7 | 54 | Chad | 0.34 | 141 |
| Moldova (h-HDI) | 3.5 | 78 | Jamaica (h-HDI) | 1.7 | 3.1 | Senegal | 0.34 | 115 |
| Lithuania (vh-HDI) | 3.4 | 28 | Nicaragua | 1.7 | 42 | Guinea-Bissau | 0.33 | 361 |
| Hungary (vh-HDI) | 3.3 | 4.5 | Argentina(vh-HDI) | 1.6 | 28 | Bangladesh | 0.32 | 221 |
| Poland (vh-HDI) | 3.3 | 9.4 | Mauritius (h-HDI) | 1.6 | 12 | Gabon (h-HDI) | 0.31 | 517 |
| Serbia (vh-HDI) | 3.3 | 14 | Romania (vh-HDI) | 1.6 | 43 | Niger | 0.31 | 82 |
| Australia (NB; vh-HDI) | 3.2 | 7.2 | Thailand (vh-HDI) | 1.6 | 144 | Namibia | 0.3 | 451 |
| Germany (NB; vh-HDI) | 3.2 | 5.3 | Tunisia (h-HDI) | 1.6 | 36 | Nepal | 0.3 | 230 |
| Portugal (NB; vh-HDI) | 3.2 | 16 | Bosnia and Herzegovina (h-HDI) | 1.5 | 26 | Sudan | 0.3 | 62 |
| Russia (vh-HDI) | 3.1 | 48 | Cyprus (NB; vh-HDI) | 1.5 | 3.2 | Angola | 0.26 | 329 |
| Albania (h-HDI) | 3 | 16 | Qatar (vh-HDI) | 1.5 | 36 | Sierra Leone | 0.26 | 292 |
| Italy (NB; vh-HDI) | 3 | 4.2 | Yemen | 1.5 | 48 | Cameroon | 0.22 | 171 |
| Turkey (vh-HDI) | 3 | 16 | Dominican Republic (h-HDI) | 1.4 | 41 | Burkina Faso | 0.17 | 46 |
| United Kingdom (NB; vh-HDI) | 3 | 7 | Kazakhstan (vh-HDI) | 1.4 | 66 | Madagascar | 0.17 | 233 |
| Uruguay (vh-HDI) | 3 | 32 | Saudi Arabia (vh-HDI) | 1.4 | 7.8 | Democratic Republic of Congo | 0.16 | 319 |
| Armenia (h-HDI) | 2.9 | 25 | Cambodia | 1.3 | 280 | Togo | 0.16 | 35 |
| Austria (NB; vh-HDI) | 2.9 | 4.9 | Honduras | 1.3 | 30 | Central African Republic | 0.14 | 540 |
| Ireland (NB; vh-HDI) | 2.9 | 5.3 | Kyrgyzstan | 1.3 | 108 | Guinea | 0.14 | 175 |
| The Netherlands (NB; vh-HDI) | 2.9 | 4.1 | Laos | 1.3 | 149 | Liberia | 0.14 | 308 |
| Belgium (NB; vh-HDI) | 2.8 | 7.7 | Philippines (h-HDI) | 1.3 | 533 | Cote d'Ivoire | 0.1 | 132 |
| Spain (NB; vh-HDI) | 2.8 | 7.2 | Turkmenistan (h-HDI) | 1.2 | 46 | Tanzania | 0.08 | 222 |
| Chile (vh-HDI) | 2.7 | 14 | Uzbekistan (h-HDI) | 1.2 | 65 | Bahamas (NB; vh-HDI) | 0 | 8.8 |
| Czechia (NB; vh-HDI) | 2.7 | 3.9 | India | 1.1 | 204 | Barbados (h-HDI) | 0 | 2.5 |
| France (**NB**; vh-HDI) | 2.7 | 8.3 | Paraguay (h-HDI) | 1.1 | 45 | Belize | 0 | 27 |
| Malaysia (vh-HDI) | 2.7 | 90 | Vietnam (h-HDI) | 1.1 | 171 | Benin | 0 | 54 |
| Mexico (h-HDI) | 2.7 | 23 | Afghanistan | 1 | 189 | Bhutan | 0 | 164 |
| New Zealand (**NB**; vh-HDI) | 2.7 | 7.2 | Kuwait (vh-HDI) | 1 | 18 | Brunei (vh-HDI) | 0 | 82 |
| Norway (**NB**; vh-HDI) | 2.7 | 3.1 | Panama (vh-HDI) | 1 | 33 | Comoros | 0 | 35 |
| Palestine (h-HDI) | 2.7 | 0.65 | Sri Lanka (h-HDI) | 1 | 63 | Congo | 0 | 372 |
| Denmark (**NB**; vh-HDI) | 2.6 | 4.1 | North Korea | 0.97 | 513 | Djibouti | 0 | 220 |
| Ukraine (h-HDI) | 2.6 | 74 | Tajikistan | 0.96 | 80 | Equatorial Guinea | 0 | 275 |
| Japan (vh-HDI) | 2.5 | 12 | Bahrain (vh-HDI) | 0.93 | 15 | Fiji (h-HDI) | 0 | 66 |
| Jordan (h-HDI) | 2.5 | 4.6 | Ghana | 0.89 | 140 | Gambia | 0 | 153 |
| Sweden (**NB**; vh-HDI) | 2.5 | 3.5 | Azerbaijan (h-HDI) | 0.85 | 58 | *Guam (vh-HDI) | 0 | 39 |
| Algeria (h-HDI) | 2.4 | 58 | Nigeria | 0.83 | 219 | Guyana (h-HDI) | 0 | 78 |
| *Puerto Rico (**NB**; vh-HDI) | 2.4 | 0.95 | Zimbabwe | 0.81 | 181 | Haiti | 0 | 164 |
| Switzerland (**NB**; vh-HDI) | 2.4 | 4.7 | Indonesia (h-HDI) | 0.79 | 301 | Iceland (**NB**; vh-HDI) | 0 | 3.8 |
| Costa Rica (vh-HDI) | 2.3 | 9.7 | Zambia | 0.75 | 319 | Luxembourg (**NB**; vh-HDI) | 0 | 5.8 |
| Cuba (h-HDI) | 2.3 | 6.3 | Botswana (h-HDI) | 0.74 | 230 | Maldives (h-HDI) | 0 | 37 |
| Egypt (h-HDI) | 2.3 | 11 | Eswatini | 0.7 | 342 | Malta (**NB**; vh-HDI) | 0 | 31 |
| Finland (**NB**; vh-HDI) | 2.3 | 3.6 | Kenya | 0.7 | 250 | Papua New Guinea | 0 | 418 |
| Lebanon (**NB**; h-HDI) | 2.3 | 13 | South Africa (h-HDI) | 0.68 | 562 | Saint Lucia (h-HDI) | 0 | 2.2 |
| South Korea (vh-HDI) | 2.3 | 48 | Bolivia | 0.67 | 103 | Samoa (h-HDI) | 0 | 7 |
| Trinidad and Tobago (**NB**; vh-HDI) | 2.3 | 17 | Pakistan | 0.65 | 255 | Sao Tome and Principe | 0 | 115 |
| Brazil (h-HDI) | 2.2 | 45 | Somalia | 0.64 | 254 | Solomon Islands | 0 | 58 |
| Bulgaria (vh-HDI) | 2.2 | 19 | Ethiopia | 0.61 | 129 | Vanuatu | 0 | 37 |
| Ecuador (h-HDI) | 2.2 | 44 | Timor | 0.56 | 473 |  |  |  |
| **Sub-Average** | **3.04** | **17.46** |  | **1.32** | **125.58** |  | **0.18** | **171.09** |
| **Sub-STDEV** | **0.67** | **19.93** |  | **0.48** | **139.41** |  | **0.19** | **152.30** |
| **AVERAGE (ALL COUNTRIES)** | **1.52** | **104.34** |  |  |  |  |  |  |
| **STDEV (ALL COUNTRIES)** | **1.27** | **135.29** |  |  |  |  |  |  |

CORRELATION ANALYSIS
| **Countries** |  | **BCNS Cancer Inc. in 0-4Y-olds vs TB Inc. per 100k** |  |
| --- | --- | --- | --- |
|  |  | Affected and Not affected Countries (BCNS Inc.: 0-5.4) | Affected Countries only (BCNS Inc.: >0 - 5.4) |
| **All** |  | **r(179): -0.4973; P = < 0.0001** | **r(154): -0.6085; P = < 0.0001** |
| **All: NB or No-BCG Only** |  | r(33): -0.2179; P = 0.2232 | r(29): +0.0362; P = 0.8522 |
| **All: BCG Only** |  | **r(146): -0.4581; P = < 0.0001** | **r(125): -0.5595; P = < 0.0001** |
| **HDI Sub-Group** | **vh- & h-HDI: All** | **vh-&h-HDI: r(108): -0.3428; P = < 0.0003**<br>**vh-HDI: r(64): -0.3040; P = 0.0146**<br>**h-HDI: r(44): -0.3450; P = 0.0218** | **vh-&h-HDI: r(96): -0.4995; P = < 0.0001**<br>**vh-HDI: r(58): -0.3313; P = 0.0111**<br>**h-HDI: r(38): -0.5885; P = 0.0001** |
|  | **vh- & h-HDI: NB only** | **vh-&h-HDI: r(33): -0.2179; P = 0.2232**<br>**vh-HDI: r(31): -0.3903; P = 0.0299** | **vh-&h-HDI: r(29): +0.0362; P = 0.8522**<br>**vh-HDI: r(27): -0.1462; P = 0.4670** |
|  | **vh- & h-HDI: BCG only** | **vh-&h-HDI: r(75): -0.3412; P = < 0.0027**<br>vh-HDI: r(33): -0.3427; P = 0.0509 | **vh-&h-HDI: r(67): -0.4825; P = < 0.0001**<br>vh-HDI: r(31): -0.2668; P = 0.1467 |
|  |  | **h-HDI: $r(42)$: -0.3359; $P$ = 0.0297** | **h-HDI: $r(36)$: -0.5993; $P$ = 0.0001** |
| **Note:** The values shown are rounded off to the indicated decimal places; STDEV: standard deviation; Exclusion of outliers in No-BCG and BCG Countries, i.e., Incidence rate '0' and Maximum '5.4', respectively or only consideration of countries with BCNS Inc.: >0 to 4.3, did not adversely affect the correlation for groups 'All countries' [$r(153)$: -0.6151; $P$ = <0.0001] or 'BCG Countries' [$r(124)$: -0.5679; $P$ = <0.0001].<br><sup>a</sup>Human Development Index (HDI) 2020: A composite index used by United Nations Development Program (UNDP) to indicate overall human development in three basic dimension, i.e., "a long healthy life, knowledge base and a decent standard of living"[50]. Countries with HDI >0.8 and 0.7-0.799 are considered as very high (vh-) and high (h-) HDI countries, respectively. *Not listed by UNDP. Some countries were shown as vh- and h-HDI by UNDP in 2020 but not by IACR [34] were considered as such for correlation analysis.<br><sup>b</sup>[37]. Age-standardized rate (ASR) estimates for BCNS cancer in 0-4Y-old (2020) children are from the GLOBOCAN 2020 study, International Agency for Research on Cancer (IARC), World Health Organization. The sources and methodology employed for the global cancer incidence estimates for GLOBOCAN 2020 are described at the Global Cancer Observatory (GCO) website gco.iarc.fr [Available at https://gco.iarc.fr/today/home; Last accessed 15 December 2022].<br><sup>c</sup>[51] The estimates of TB incidence for countries in 2020 from Global Tuberculosis Programme: WHO TB burden estimates. Available at https://www.who.int/teams/global-tuberculosis-programme/data [51][Last accessed 05 Feb 2023] |  |  |  |

### BCG Countries Frequently Have Lower Incidence Of Early Childhood BCNS Cancer

Globally, the distribution of updated age-standardized incidence rates (ASR) of BCNS cancer for the recent year 2020 [37] varies widely from 0 to 5.4 per 100,000 (100k) in 0-4Y-olds (Table 1). Nevertheless, the widely mandated BCG vaccination seems associated with reduced childhood BCNS cancer incidences in 0-4Y-olds (Figure 1A). The countries of Europe and North America with no neonatal BCG vaccination policy in place (‘No-BCG’ or ‘NB’) are some of the worst affected countries, while incidence rates in countries with BCG vaccination in place (BCG countries) do have some similar high incidence rate countries, but the majority seem to cluster towards lower incidence rates. The incidences in No-BCG and BCG countries significantly differ (Mean: 2.64 *vs* 1.26; Median: 2.8 *vs* 0.985; IQR; 2.4–3.2 *vs* 0.31–2.0; *P* = <0.0001 (two-tailed)). The high to low incidence rates of BCNS cancer among countries also display an interesting association with countries’ income brackets [49]. The higher income brackets have higher average incidence rates than the lower income brackets (Figure 1A: right side Y axis). The previous observations made in small-scale studies performed in different countries that identified the socioeconomic position (SEP) of families as a potential risk factor positively associated with childhood BCNS cancer incidences [52,53], seem to be applicable globally. However, the occurrence of high incidence rates (ASR) in certain BCG vaccinating countries of Europe [notably, Montenegro (5.4), Estonia (4.3), Croatia (4.3), North Macedonia (3.6), Republic of Moldova (3.5), Lithuania (3.4), Serbia, Poland (3.3)] along with wide variation in incidence rates among BCG vaccinating countries 0 – 5.4, casts doubt on the ability of single BCG shots alone to protect young children from BCNS cancers, as could have been the case in some of the past studies reported in [15,19,20,22] that failed to find an association between BCG vaccination and cancer incidence.

**Figure 1.**
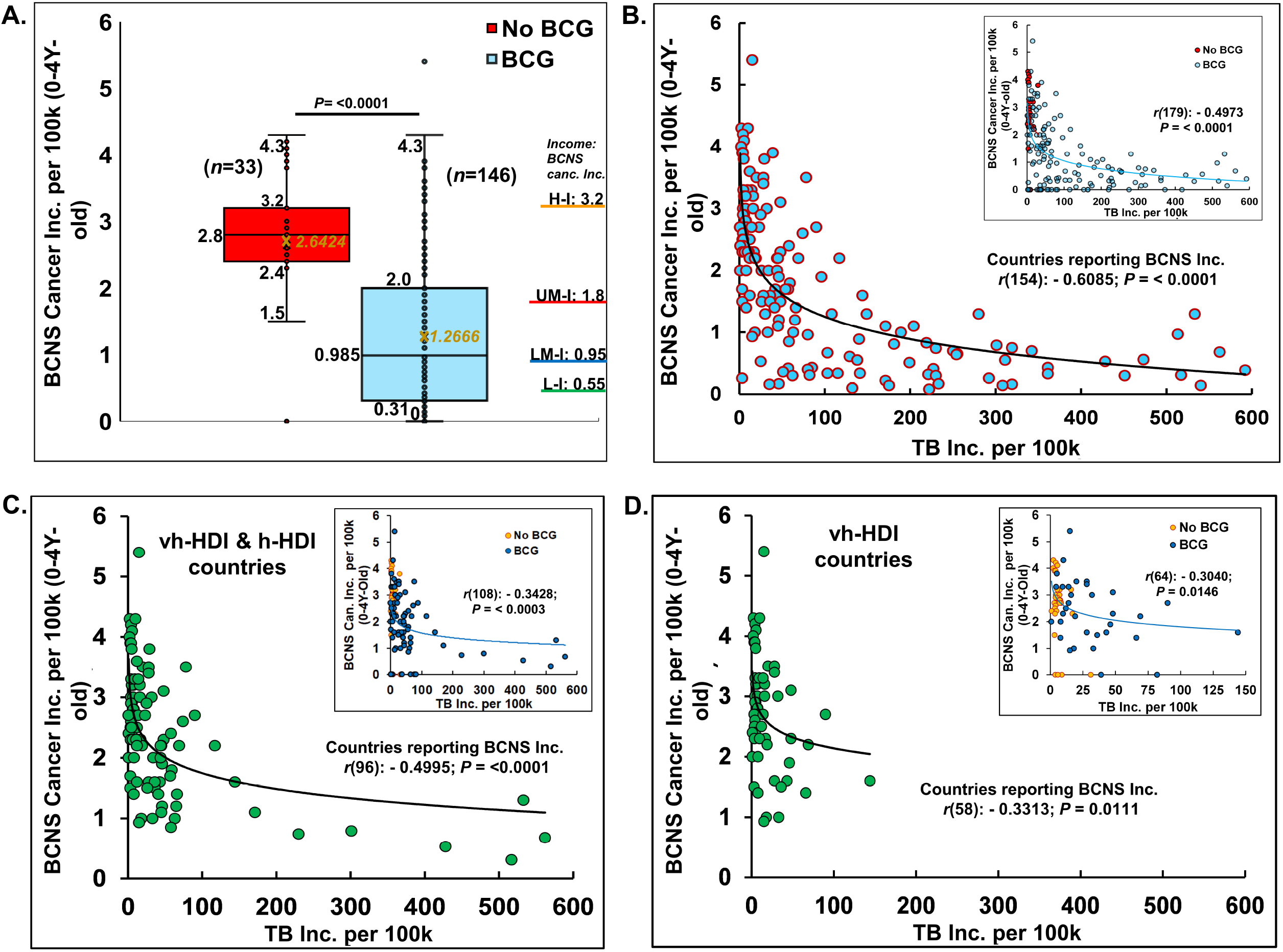
BCG vaccination and *M. tuberculosis* complex exposure negatively associated with BCNS cancer incidences in 0-4Y-old children. Globally, the distribution of age-standardized BCNS cancer incidence rates (ASR) among countries (n=179) are shown as Box and whisker plot **(A)**. BCG-vaccinated children had significantly reduced incidences (t-test: two-tailed, equal/unequal variance) than the non-vaccinated (No BCG). The average incidence rate seems directly proportional to income levels [H-I: High ($13,205 or more); UM-I: Upper Middle (between $4,256 and $13,205); LM-I: Lower Middle ($1,086 and $4,255); L-I: Low income (≤$1,086) as defined by World Bank, 2023 [49] (For CI refer to [37]). The probability of natural boosting (exposure to the *M. tuberculosis* complex) is associated with lower BCNS cancer incidence in the affected countries/territories: All **(B)**; vh-HDI and h-HDI **(C)** and vh-HDI countries **(D)**. Inset has countries color-coded for BCG and No BCG groups. The trend lines are drawn to guide the eye. *Note*: Outliers presence in No-BCG and BCG groups (A). Most of the high BCNS incidence displaying countries of the BCG group including the outlier are clustered in low background *M. tuberculosis complex* exposure regions with TB incidence rate <100 per 100k (B,C) for the year 2020.

### Globally, Higher Natural “Boosting” Potential Negatively Associated With Pediatric BCNS Cancer Incidences In Countries

A missing explanatory variable for the observed reduced BCNS cancer incidences in BCG countries could be the ‘boosting’ stimulus (also priming for No-BCG countries) arising from natural exposure to boosting events in the form of *M. tuberculosis* complex exposure specific to populations. It may be pertinent here to reiterate for the scientists from other backgrounds that *M. tuberculosis* complex exposure does not invariably cause TB but rather only sustained long-term exposure of susceptible individuals, which constitutes a small fraction of the population – variously estimated to be <1-5%, is supposed to result in clinical TB [REFs. 33,54]. Assuming the BCG vaccination could be playing a role in the BCNS cancer incidences in young children, the higher incidences in some countries could be envisioned to result from the inability of a single BCG inoculation to sufficiently activate the immune system in the absence of proper, timely boosting. Revaccination has been indicated to enhance the non-specific protective effects of BCG in early childhood [REFs. 3,55,56]. The neonates in countries with higher TB prevalence or incidence rates [51] could be more frequently exposed to natural boosting events than those born in low TB incidence countries.

Surprisingly, when we look at the covariation of BCNS cancer incidence rates [37] with countries’ TB incidence rates [51] they are found to be significantly negatively correlated even when completely disregarding their BCG vaccination policy (Table 1). This correlation further improves on consideration of the countries reporting childhood BCNS cancer incidences [Table 1, Affected countries: *r*(154): - 0.6085, *P* = <0.0001; All countries (affected + non-affected) *r*(179): - 0.4973; *P*= <0.0001]. The consideration of only BCG countries slightly decreases the correlation that could be potentially indicative of the important role of exposure to the *M. tuberculosis* complex in No-BCG countries as well. Looking at the distribution of BCNS cancer incidences, it seems to be decreasing exponentially with the increase in TB incidence rates of the countries (*i*.*e*., increased probability of natural exposure: priming and boosting). The seeming outlier of the BCG vaccination group that reported BCNS cancer incidences of 5.4 (i.e., Montenegro) as well as other high-incidence reporting countries happened to be some of the least TB incidence (*M. tuberculosis* complex exposure) countries of the No-BCG and BCG group of countries (Figure 1B). The increase in TB incidence rates up to 100 per 100,000 (100k), is seemingly associated with a four-fold reduction in BCNS cancer incidences among 0-4Y-olds, which further goes down by two-fold on an increase in TB incidence to 200 per 100k. The same trend follows till the highest TB incidence rate of about 600 per 100k, potentially indicating TB incidence rates in populations as a risk factor inversely correlated with early childhood BCNS cancer incidences.

### In Very High- and High-Human Development Index (HDI) Countries, Childhood BCNS Cancer Incidences Negatively Associated With the Natural “Boosting” Possibility

The reporting of childhood BCNS cancer incidences across countries has not been uniform. The projected estimates could be more skewed in low HDI countries because of the possible lack of education, medical infrastructure, diagnostics, affordability, and reporting. Historical estimates for many countries display fluctuations in 3–8-year periods with extreme variations in sex-specific incidences, potentially indicative of more imprecise estimates, and underline a need for better data reporting and collation than current [37]. Thus, consideration of the latest BCNS cancer incidence estimates in very high HDI (vh-HDI) and high HDI (h-HDI) countries may be expected to more reliably illustrate the association, if any. Remarkably, the BCNS cancer incidence among 0-4Y-olds in vh-HDI and h-HDI countries during 2020 was also found to be significantly but negatively correlated with their TB incidence rates [Figure 1C-D; Table 1, Affected vh-& h-HDI: *r*(96): −0.4995; *P* = < 0.0001; All (affected + non-affected) vh-& h-HDI: *r*(108): −0.3428, *P* = < 0.0003). For the associations in various subgroups, refer to Table 1. The negative association between childhood BCNS cancer incidences and exposure to the *M. tuberculosis* complex remained significant for vh-HDI and h-HDI countries separately as well (Table 1 subgroups).

## DISCUSSION

The early childhood BCNS cancer incidence in countries appears to be negatively associated with the prevailing possibility of exposure to the *M. tuberculosis* complex. The observations presented could be construed as indicative of non-exposure to *M. tuberculosis* complex or “boosting” as an unidentified risk factor for childhood BCNS cancer, possibly functionally related to the “Hygiene Hypothesis” and associated trained immunity. The boosting event(s) could be necessary for supposed priming or activation (immunomodulation), and reprogramming of the metabolism for desired outcomes. We suspect the current regimen of single neonatal BCG inoculation may not be sufficient to cause desired immune activations in environments lacking boosting opportunities. Possibly, multiple shot regimens, as used to be given earlier in many countries [39] or suggested for enhancing nonspecific effects [55,56] may be necessary for providing the required stimulus and boosting to stem the steady annual rise of early childhood BCNS cancer incidences globally [37]. However, since a large majority of early childhood BCNS cancers are supposedly embryonic developmental remnants, the likelihood of the mother’s immune system getting modulated by exposure to BCG and/or *M. tuberculosis* complex itself and in effect modulating and moderating that of the embryo (*in utero*, including endocrine) for ‘normal’ development remains a possibility [20,28]. We would be inclined to suggest the pouring and pooling of resources for a thorough evaluation of BCG’s potential in reducing childhood BCNS cancer incidences. With BCNS cancers in children on the rise, carefully planned, appropriately controlled, multicentric clinical trials preferably coordinated and managed by WHO be performed in high-incidence countries, both No-BCG and BCG countries to reliably evaluate the qualitative and quantitative contributions of neonatal BCG inoculation(s), boosting, and the mother’s immune system modulation. The countries with zero incidence rates could provide additional insights in terms of etiological agents as well as help identify unknown risk factors specific to countries that are responsible for huge differences in the BCNS cancer incidence rate even among socially similar vh-HDI countries (*e*.*g*., Iceland, Luxembourg *vs*. Germany, Ireland, Greece, Montenegro, etc.). If immunomodulation has a role in the increased incidence of BCNS cancers in childhood, the recent change in schedule and policy from multiple to single to no BCG inoculations could be costing us >10-fold higher incidence and mortality.

The inherent limitations of the cancer incidences dataset arising from data availability and methodology employed [57] should also be considered when planning for exploratory evaluation studies. As BCNS cancer incidences and reporting globally remain fragmentary [37,57], the studies, whether prospective or registry-based retrospective, should wherever possible be performed in high-incidence but low TB prevalence countries, carefully controlling for age, trained immunity status, mothers’ immunization, *etc*. to arrive at meaningful conclusions about its preventive interventional potential. Consideration of existing trends of incidence in previous relevant years [37] as a baseline could provide additional pointers to help reliably estimate the impact of specific interventions in clinical trials.

## CONCLUSION

BCG vaccinations coupled with timely natural “boosting” could be causally associated with a reduction in childhood BCNS cancer incidences. Future research may be directed at evaluating the same in carefully controlled trials and identifying country-specific differences and modifiers. It is needed to improve our understanding of the role of BCG vaccination and the mechanistic details of the potential protective immune augmentation process. It may help devise ways to stem the steady rise of early childhood cancer incidences with similar components. Additionally, the findings could indicate inoculation regimens that are better suited for lowering early childhood cancer incidences. The revised multiple BCG vaccinations, as they used to be, may have to be reintroduced globally to reduce childhood cancer incidences. The countries with higher incidence rates (ASR per 100k), *e*.*g*., Montenegro (5.4), Estonia (4.3), USA (4.3), Greece (4.2), Canada (4.1), Israel (4), Slovakia (3.9), Slovenia (3.9), etc., are going to gain the most in terms of morbidity and mortality reduction. However, a lot needs to be done to reliably ascertain the potential benefits of BCG vaccination in reducing cancer incidence, or the lack thereof, before this could be again reintroduced or written off.

## MATERIALS AND METHODS

The estimates of cancer incidence rates in different age groups in individual countries are from GLOBOCAN 2020, International Agency for Research on Cancer, WHO [37]. The BCG vaccination policy information is from the World Atlas of BCG Policies and Practices, 3rd Edition [39]. The categorization of countries in different income groups is from the World Bank Group, 2023 [49]. The countries’ categorization as very high- and high-Human Development Index in 2020 is primarily from Human Development Report 2021/2022 by United Nations Development Programme [50]. The estimates of Tuberculosis incidence in 2020 are from WHO, Global Tuberculosis Programme [51]. Basic statistical analysis is performed using Excel 2019. The *P*-value <0.05 is reported as significant.

## Data Availability

All data produced in the present work are contained in the manuscript and available online

## Acknowledgments

The laboratory of SS is supported by a seed grant from IoE, Banaras Hindu University. The work presented is unfunded.

## Author Contributions

SS conceived the idea, designed the research, analyzed the data, and wrote the paper. RKS and AD wrote the paper.

## Compliance with Ethical Standards

### Data Availability Statement

All pertinent data is available in the public domain and is included in the manuscript and the associated supplementary file. The raw data generated as a part of the analysis is also available on request.

### Ethics statement

The study complies with ethical requirements and standards. No identifiable human data is used.

## Figure Legends

**Supplementary Figure 1. Estimated Annual Percentage Change of Brain and Central Nervous System Cancer Incidence ASR per 100**,**000 in 0-4Y-olds**. Majority of No-BCG and BCG countries/territories (n=44) display positive annual percentage change in combined incidences for sexes in 0-4Y-olds (last 15 years data). Bars show CI (Available at https://gco.iarc.fr/overtime/en/dataviz/eapc?populations=38000_3600_7600_10000_11200_12400_15200_15600_17000_19100_18800_20300_20800_21800_25000_27600_23300_35200_35600_37200_37600_39200_41000_41400_42800_44000_47000_52800_55400_57800_6 1600_70300_70500_72400_75200_75600_76400_79200_80000_80400_82630_82610_84000_82620&sexes=1_2&multiple_populations=1&years=2018&cancers=23&types=0&key=asr&ag e_end=0&group_cancers=0&multiple_cancers=0&mode=population&age_start=0&group_year s=0&eapc_span=15&ul=1). Note: The historical estimates remain weak. The observed sex-specific incidence variation in the past for specific years or over time could be indicative of reporting issues. (Available at https://gco.iarc.fr/overtime/en/dataviz/trends?populations=38000&sexes=1_2&multiple_po pulations=1&years=1943_2018&cancers=23&types=0&key=asr&age_end=0&group_cancers= 0&multiple_cancers=0&mode=population&age_start=0&group_years=0).

## REFERENCES

1. UNICEF. Bacillus Calmette-Guérin (BCG) supply and demand update; 2019. [Available at https://www.unicef.org/supply/reports/bacillus-calmette-gu%C3%A9rin-bcg-supply-and-demand-update]

2. Trunz BB, Fine P, Dye C. Effect of BCG vaccination on childhood tuberculous meningitis and miliary tuberculosis worldwide: a meta-analysis and assessment of cost-effectiveness. Lancet. 2006 Apr 8;367(9517):1173–80. doi: 10.1016/S0140-6736(06)68507-3.

3. Trunk G, Davidović M, Bohlius J. Non-Specific Effects of Bacillus Calmette-Guérin: A Systematic Review and Meta-Analysis of Randomized Controlled Trials. Vaccines (Basel). 2023 Jan 4;11(1):121. doi: 10.3390/vaccines11010121.

4. Biering-Sørensen S, Aaby P, Lund N et al. Early BCG and neonatal mortality among low-birth-weight infants: A randomised controlled trial. Clin Inf Dis 2017; 65: 1183–90.

5. Higgins JPT, Soares-Weiser K, Lopez-Lopez J A et al. Association of BCG, DTP, and measles containing vaccines with childhood mortality: systematic review. BMJ 2016; 355: i5170

6. Aaby P, Roth A, Ravn H, Napirna BM, Rodrigues A, Lisse IM, Stensballe L, Diness BR, Lausch KR, Lund N, Biering-Sørensen S, Whittle H, Benn CS. Randomized trial of BCG vaccination at birth to low-birth-weight children: beneficial nonspecific effects in the neonatal period? J Infect Dis. 2011 Jul 15;204(2):245–52. doi: 10.1093/infdis/jir240.

7. Nankabirwa, V., Tumwine, J.K., Mugaba, P.M. et al. Child survival and BCG vaccination: a community based prospective cohort study in Uganda. BMC Public Health 15, 175 (2015). https://doi.org/10.1186/s12889-015-1497-8

8. Brook B, Harbeson DJ, Shannon CP, Cai B, He D, Ben-Othman R, et al. BCG vaccination-induced emergency granulopoiesis provides rapid protection from neonatal sepsis. Sci Transl Med. 2020 May 6;12(542):eaax4517. doi: 10.1126/scitranslmed.aax4517.

9. Netea MG, Joosten LA, Latz E, Mills KH, Natoli G, Stunnenberg HG, et al. Trained immunity: A program of innate immune memory in health and disease. Science. 2016 Apr 22;352(6284):aaf1098. doi: 10.1126/science.aaf1098.

10. Cvián C, Fernández-Fierro A, Retamal-Díaz A, Díaz FE, Vasquez AE, Lay MK, et al. BCG-Induced Cross-Protection and Development of Trained Immunity: Implication for Vaccine Design. Front Immunol. 2019 Nov 29;10:2806. doi: 10.3389/fimmu.2019.02806.

11. Benn CS, Netea MG, Selin LK, Aaby P. A small jab - a big effect: nonspecific immunomodulation by vaccines. Trends Immunol. 2013 Sep;34(9):431–9. doi: 10.1016/j.it.2013.04.004.

12. Kleinnijenhuis J, Quintin J, Preijers F, Joosten LA, Ifrim DC, Saeed S, et al. Bacille Calmette-Guerin induces NOD2-dependent nonspecific protection from reinfection via epigenetic reprogramming of monocytes. Proc Natl Acad Sci U S A. 2012 Oct 23;109(43):17537–42. doi: 10.1073/pnas.1202870109.

13. Arts RJ, Joosten LA, Netea MG. Immunometabolic circuits in trained immunity. Semin Immunol. 2016 Oct;28(5):425–430. doi: 10.1016/j.smim.2016.09.002.

14. Cheng SC, Quintin J, Cramer RA, Shepardson KM, Saeed S, Kumar V, et al. mTOR- and HIF-1α-mediated aerobic glycolysis as metabolic basis for trained immunity. Science. 2014 Sep 26;345(6204):1250684. doi: 10.1126/science.1250684.

15. Singh AK, Netea MG, Bishai WR. BCG turns 100: its nontraditional uses against viruses, cancer, and immunologic diseases. J Clin Invest. 2021 Jun 1;131(11):e148291. doi: 10.1172/JCI148291.

16. Saeed S, Quintin J, Kerstens HHD, Rao NA, Aghajanirefah A, Matarese F, et al. Epigenetic programming of monocyte-to-macrophage differentiation and trained innate immunity. Science. (2014) 345:1251086. doi: 10.1126/science.1251086

17. Greaves M. A causal mechanism for childhood acute lymphoblastic leukaemia. Nat Rev Cancer. 2018 Aug;18(8):471–484. doi: 10.1038/s41568-018-0015-6.

18. Rosenthal SR. Cancer precursors and their control by BCG. Dev Biol Stand. 1986;58 (Pt A):401–16.

19. Grange JM, Stanford JL. BCG vaccination and cancer. Tubercle. 1990 Mar;71(1):61–4. doi: 10.1016/0041-3879(90)90063-e.

20. Grange JM, Stanford JL, Stanford CA, Kölmel KF. Vaccination strategies to reduce the risk of leukaemia and melanoma. J R Soc Med. 2003 Aug;96(8):389–92. doi: 10.1258/jrsm.96.8.389

21. Angelidou A, Pittet LF, Faustman D, Curtis N, Levy O. BCG vaccine’s off-target effects on allergic, inflammatory, and autoimmune diseases: Worth another shot? J Allergy Clin Immunol. 2022 Jan;149(1):51–54. doi: 10.1016/j.jaci.2021.09.034.

22. Marron M, Brackmann LK, Kuhse P, Christianson L, Langner I, Haug U, Ahrens W. Vaccination and the Risk of Childhood Cancer-A Systematic Review and Meta-Analysis. Front Oncol. 2021 Jan 2;10:610843. doi: 10.3389/fonc.2020.610843.

23. Davignon L, Robillard P, Lemonde P, Frappier A. B.C.G. vaccination and leukemia mortality. Lancet. 1970 Sep 26;2(7674):638. doi: 10.1016/s0140-6736(70)91402-9.

24. Rosenthal SR, Crispen RG, Thorne MG, Piekarski N, Raisys N, Rettig PG. BCG vaccination and leukemia mortality. JAMA. 1972 Dec 18;222(12):1543–4.

25. Snider DE, Comstock GW, Martinez I, Caras GJ. Efficacy of BCG vaccination in prevention of cancer: an update. J Natl Cancer Inst. 1978 Apr;60(4):785–8. doi: 10.1093/jnci/60.4.785.

26. Nathanson L. Use of BCG in the treatment of human neoplasms: a review. Semin Oncol. 1974;1(4):337–350.

27. Bast RC Jr, Zbar B, Borsos T, Rapp HJ. BCG and cancer. N Engl J Med. 1974 Jun 27;290(26):1458–69. doi: 10.1056/NEJM197406272902605.

28. Hauer J, Fischer U, Borkhardt A. Toward prevention of childhood ALL by early-life immune training. Blood. 2021 Oct 21;138(16):1412–1428. doi: 10.1182/blood.2020009895.

29. Angelidou A, Conti MG, Diray-Arce J, Benn CS, Shann F, Netea MG, et al. Licensed Bacille Calmette-Guérin (BCG) formulations differ markedly in bacterial viability, RNA content and innate immune activation. Vaccine. 2020 Feb 24;38(9):2229–2240. doi: 10.1016/j.vaccine.2019.11.060.

30. Miyasaka M. Is BCG vaccination causally related to reduced COVID-19 mortality? EMBO Mol Med. 2020 Jun 8;12(6):e12661. doi: 10.15252/emmm.202012661.

31. Rakshit S, Ahmed A, Adiga V, Sundararaj BK, Sahoo PN, Kenneth Jet al. BCG revaccination boosts adaptive polyfunctional Th1/Th17 and innate effectors in IGRA+ and IGRA–Indian adults. JCI Insight.2019; 4 (24):e130540. https://doi.org/10.1172/jci.insight.130540.

32. Menzies D. Interpretation of repeated tuberculin tests. Boosting, conversion, and reversion. Am J Respir Crit Care Med. 1999 Jan;159(1):15–21. doi: 10.1164/ajrccm.159.1.9801120.

33. Singh S, Maurya RP, Singh RK (2020) “Trained immunity” from Mycobacterium spp. exposure or BCG vaccination and COVID-19 outcomes. PLoS Pathog 16(10): e1008969. https://doi.org/10.1371/journal.ppat.1008969

34. Strachan DP. Hay fever, hygiene, and household size. BMJ (Clinical research ed). 1989;299(6710):1259–60. https://doi.org/10.1136/bmj.299.6710.1259.

35. Stanford JL, Stanford CA, Grange JM. Environmental echoes. Science Progr 2001;84(Pt 2):105–24. doi: 10.3184/003685001783239014.

36. Grange JM, Stanford JL, Stanford CA. Campbell De Morgan’s ‘Observations on cancer’, and their relevance today. J R Soc Med 2002;95:296–9. doi:10.1177/014107680209500609.

37. GCO. WHO GLOBOCAN 2020. The International Agency for Research on Cancer’ (IARC), WHO 2023 [Available at https://gco.iarc.fr/today/home; Last Accessed 05 February 2023]

38. Dara M, Acosta CD, Rusovich V, Zellweger JP, Centis R, Migliori GB; WHO EURO Childhood Task Force members. Bacille Calmette-Guérin vaccination: the current situation in Europe. Eur Respir J. 2014 Jan;43(1):24–35. doi: 10.1183/09031936.00113413.

39. Zwerling A, Behr M, Verma A, Brewer T, Menzies D, Pai M. World Atlas of BCG Policies and Practices, 3rd Edition. 2020 [Available at http://www.bcgatlas.org/index.php]

40. Adel Fahmideh M, Scheurer ME. Pediatric Brain Tumors: Descriptive Epidemiology, Risk Factors, and Future Directions. Cancer Epidemiol Biomarkers Prev. 2021 May;30(5):813–821. doi: 10.1158/1055-9965.EPI-20-1443.

41. Ostrom QT, Adel Fahmideh M, Cote DJ, Muskens IS, Schraw JM, Scheurer ME, Bondy ML. Risk factors for childhood and adult primary brain tumors. Neuro Oncol. 2019 Nov 4;21(11):1357–1375. doi: 10.1093/neuonc/noz123.

42. Ostrom QT, Francis SS, Barnholtz-Sloan JS. Epidemiology of Brain and Other CNS Tumors. Curr Neurol Neurosci Rep. 2021 Nov 24;21(12):68. doi: 10.1007/s11910-021-01152-9.

43. Nabors LB, Ammirati M, Bierman PJ, Brem H, Butowski N, Chamberlain MC, et al. Central nervous system cancers. J Natl Compr Canc Netw. 2013 Sep 1;11(9):1114–51. doi: 10.6004/jnccn.2013.0132.

44. Porter AB, Lachance DH, Johnson DR. Socioeconomic status and glioblastoma risk: a populationbased analysis. Cancer Causes Control. 2015 Feb;26(2):179–185. doi: 10.1007/s10552-014-0496-x.

45. Turner MC. Epidemiology: allergy history, IgE, and cancer. Cancer Immunol Immunother. 2012 Sep;61(9):1493–510. doi: 10.1007/s00262-011-1180-6.

46. Cui Y, Hill AW. Atopy and Specific Cancer Sites: a Review of Epidemiological Studies. Clin Rev Allergy Immunol. 2016 Dec;51(3):338–352. doi: 10.1007/s12016-016-8559-2.

47. Johnson KJ, Cullen J, Barnholtz-Sloan JS, Ostrom QT, Langer CE, Turner MC, et al. Childhood brain tumor epidemiology: a brain tumor epidemiology consortium review. Cancer Epidemiol Biomarkers Prev. 2014 Dec;23(12):2716–36. doi: 10.1158/1055-9965.EPI-14-0207.

48. Lupatsch JE, Bailey HD, Lacour B, Dufour C, Bertozzi AI, Leblond P, Faure-Conter C, Pellier I, Freycon C, Doz F, Puget S, Ducassou S, Orsi L, Clavel J. Childhood brain tumours, early infections and immune stimulation: A pooled analysis of the ESCALE and ESTELLE case-control studies (SFCE, France). Cancer Epidemiol. 2018 Feb;52:1–9. doi: 10.1016/j.canep.2017.10.015.

49. World Bank. World Bank Country and Lending Groups. The World Bank Group. 2023; [Available at https://datahelpdesk.worldbank.org/knowledgebase/articles/906519-world-bank-country-and-lending-groups ; Last accessed 05 Feb 2023]

50. UNDP. HUMAN DEVELOPMENT REPORT 2021/2022. United Nations Development Programme. United Nations. 2022. UN Plaza, New York, NY 10017 USA. ISBN. 9789211264517. https://hdr.undp.org/system/files/documents/global-report-document/hdr2021-22pdf_1.pdf

51. WHO. Global Tuberculosis Programme: WHO TB burden estimates. Available at https://www.who.int/teams/global-tuberculosis-programme/data [Last accessed 05 Feb 2023]

52. Erdmann F, Hvidtfeldt UA, Sørensen M, Raaschou-Nielsen O. Socioeconomic differences in the risk of childhood central nervous system tumors in Denmark: a nationwide registerbased casecontrol study. Cancer causes & control: CCC. 2020;31(10):915–29. https://doi.org/10.1007/s10552-020-01332-x.

53. Francis SS, Wang R, Enders C, Prado I, Wiemels JL, Ma X, Metayer C. Socioeconomic status and childhood central nervous system tumors in California. Cancer causes & control: CCC. 2021;32(1):27–39. https://doi.org/10.1007/s10552-020-01348-3.

54. WHO. Latent tuberculosis infection: updated and consolidated guidelines for programmatic management. 2018. ISBN 978-92-4-155023-9. [Available at https://apps.who.int/iris/bitstream/handle/10665/260233/9789241550239-eng.pdf;jsessionid=24D3F1097E3884502A9C1A1698348491?sequence=1]

55. Roth A, Benn CB, Ravn H et al. Effect of revaccination with BCG in early childhood on mortality: randomised trial in Guinea-Bissau. BMJ 2010; 340: c671.

56. Benn CS, Fisker AB, Whittle HC, Aaby P. Revaccination with live attenuated vaccines confer additional beneficial nonspecific effects on overall survival: a review. EBioMedicine 2016; 10: 312–7.

57. Ferlay J, Colombet M, Soerjomataram I, Mathers C, Parkin DM, Piñeros M, Znaor A, Bray F (2019). Estimating the global cancer incidence and mortality in 2020: GLOBOCAN sources and methods. Int J Cancer. 144(8):1941–1953. https://doi.org/10.1002/ijc.31937. aAvailable at https://gco.iarc.fr/today/data-sources-methods

