## Supplementary figures and images for "BCG Vaccination Policy, Natural Boosting and Pediatric Brain and CNS Tumor Incidences"

### Supplementary Figure 1

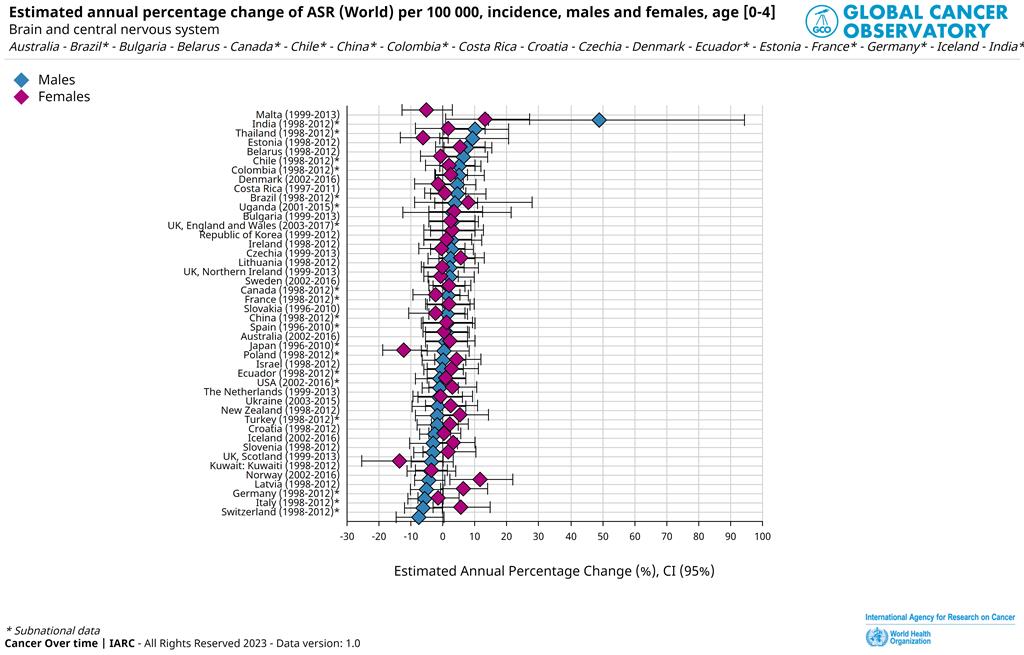
